# Clinician gaze as a supervisory signal for electrocardiogram deep learning: a matched-control proof-of-concept study

**DOI:** 10.64898/2026.11.18.26368422

**Authors:** Anurup Mukherjee, Sukhi Shergill, Jim Ang

## Abstract

**Aims:** To assess what evidence is required to attribute an apparent effect of clinician-gaze supervision on an electrocardiogram (ECG) deep-learning model to gaze itself, separating target engagement, case specificity, and diagnostic preservation.

**Methods and results:** Ten ECG stimuli interpreted by 63 readers yielded 12-lead gaze targets for nine consultants and 20 clinicians. A pretrained 23-output ECG network was fine-tuned with gaze-only loss on eight ECGs per cross-validation fold. Held-out gaze cosine improved modestly (consultants 0.858 to 0.872; all clinicians 0.884 to 0.898), but untrained training-fold mean gaze agreed better (0.929 and 0.951). Lead-occlusion concordance was weak (mean Spearman ρ 0.236). On 2,198 PTB-XL fold-10 ECGs, macro area under the receiver operating characteristic curve (AUROC) fell from 0.922 to 0.893 after gaze-only tuning; uniform and permuted controls also reduced performance. In a post hoc experiment combining PTB-XL diagnostic cross-entropy with gaze loss, macro AUROC was approximately 0.923 in diagnostic-only, real-gaze, uniform and permuted arms, but real-gaze targets were barely fitted even on training ECGs (cosine 0.858 to 0.860).

**Conclusion:** In this sparse experiment, gaze-only optimisation engaged its target without demonstrating case-specific learning beyond a population viewing prior, and matched controls showed that the resulting diagnostic deterioration was not attributable to correctly matched gaze. Joint training preserved source-domain discrimination, but because gaze targets were barely fitted, whether gaze adds diagnostic value remains untested. Claims that human viewing has shaped a model require mean-gaze, matched-null and function-preserving controls.

## Introduction

Deep-learning models can extract clinically relevant information from the 12-lead electrocardiogram across many cardiovascular conditions, but systematic reviews continue to identify limitations in external validation, robustness, calibration, and transparent interpretation [1-5]. Post-hoc visual explanations do not establish that a model uses physiologically meaningful information or reasons as a clinician would [3].

Clinician visual behaviour provides a possible source of domain knowledge. ECG interpretation involves selective inspection of leads according to rhythm, morphology and clinical context. Eye-tracking studies report differences in scan paths by expertise, while a review of 19 ECG eye-tracking studies (573 participants) identified substantial methodological heterogeneity [6-11]. Nevertheless, dwell time may represent uncertainty, difficult visual search or verification, not diagnostic importance; clinical context also changes viewing behaviour [7,10,12].

Clinician gaze has also been used as weak supervision for medical-imaging models, including radiology, pathology, endoscopy and segmentation [13-19]. However, better agreement with gaze need not imply clinically meaningful learning, and performance changes after an auxiliary loss need appropriate controls. ECG gaze research has primarily characterised interpretation behaviour rather than diagnostic-model supervision.

We asked what evidence is required before an apparent effect of gaze supervision on ECG deep learning can be attributed to clinician gaze. Three distinct claims were tested: target engagement (movement toward held-out clinician viewing), case specificity (improvement beyond a training-fold mean-gaze prior), and functional preservation (retention of diagnostic performance and attribution of any change). An apparent alignment gain or performance change alone cannot establish a gaze-specific effect.

The original hypothesis concerned target engagement alone. Training-fold mean-gaze, uniform and permuted targets, repeated derangements, lead occlusion and joint diagnostic-plus-gaze training were added after methodological review as post hoc analyses. The methodological contribution is a framework for separating the intended auxiliary target, case-specific information and clinical task performance.

## Methods

### Study design and evaluation framework

This secondary analysis of de-identified public datasets used a fixed pretrained ECG model, ten reconstructed gaze-labelled ECGs and held-out PTB-XL diagnostic evaluation (Figure 1). Table 1 identifies prespecified and post hoc components. Within each comparison, frozen starting weights, inputs, training settings and evaluation definitions were held constant, and all target permutations were fixed before inspecting their results. The subsequent joint-loss experiment used its own prespecified loss weighting and matched controls.

**Table 1.**
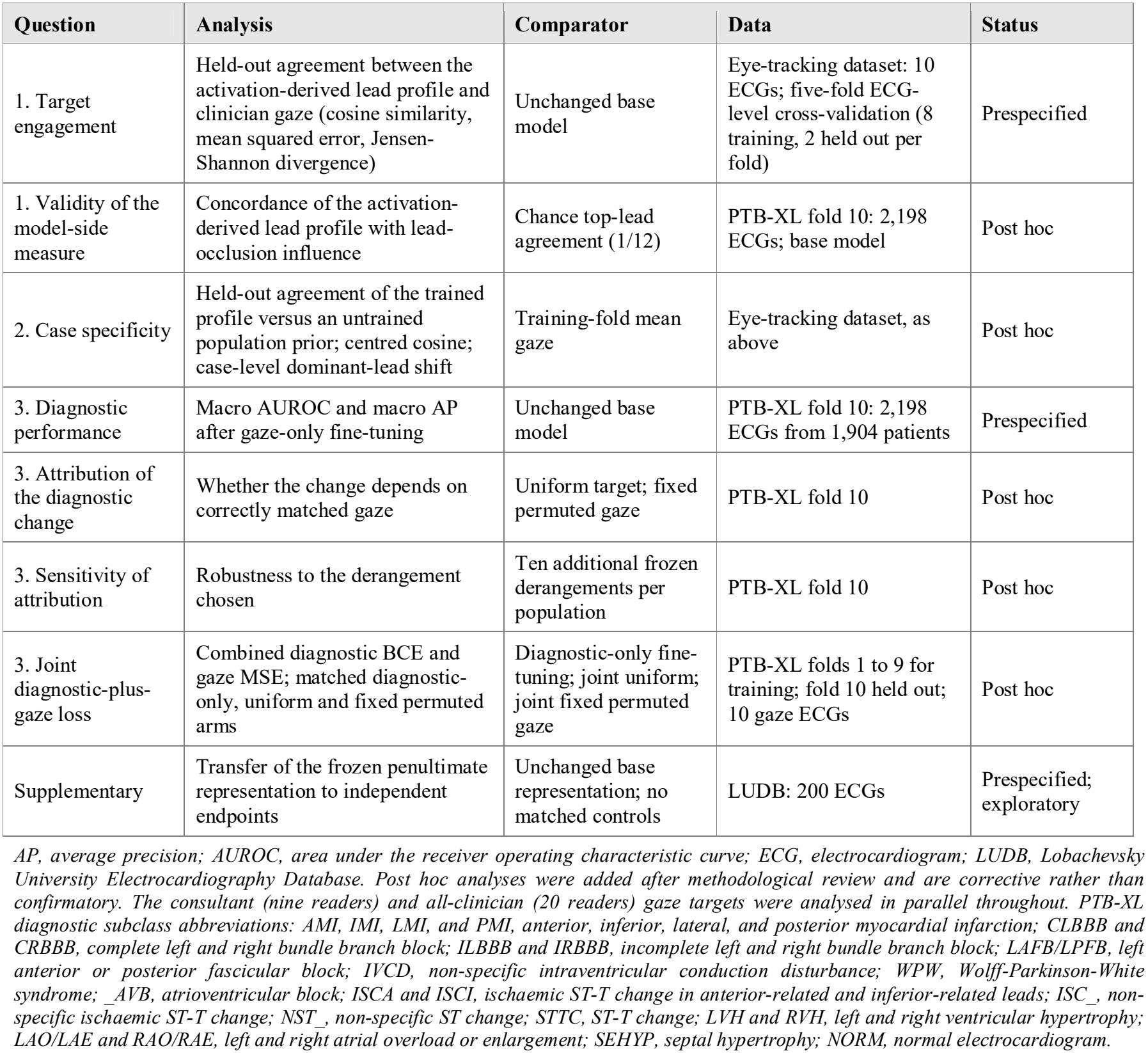
Evaluation framework: questions, analyses, comparators, data, and analysis status.

**Figure 1.**
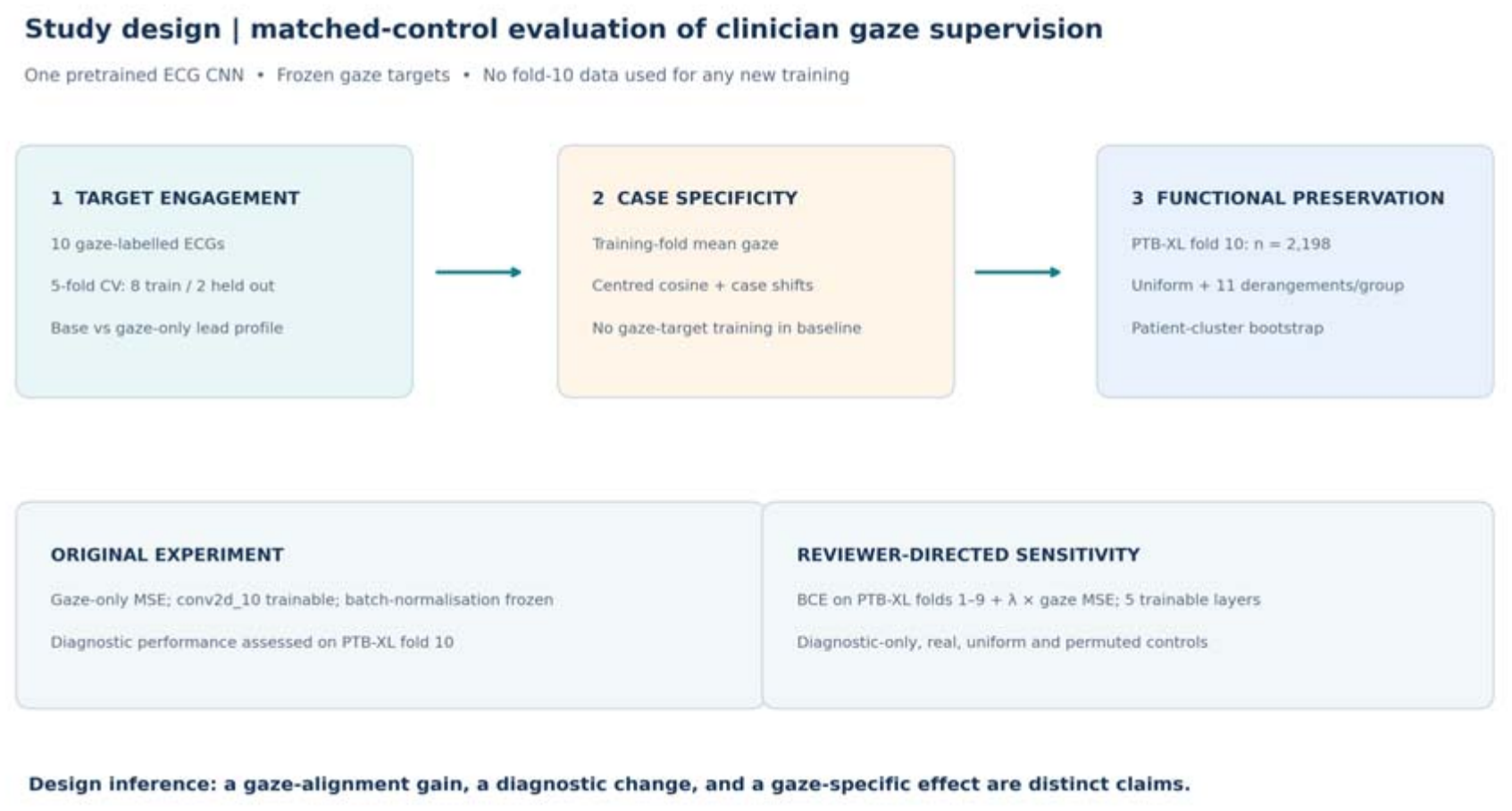
Three-question evaluation and the post hoc joint-loss experiment. The original gaze-only study tested target engagement on withheld gaze ECGs, case specificity against the training-fold mean, and diagnostic preservation against uniform and permuted auxiliary targets on PTB-XL fold 10. A separate post hoc matched experiment retained diagnostic supervision on PTB-XL folds 1 to 9 and compared diagnostic-only training against joint real-gaze, uniform and fixed-permutation controls; fold 10 remained held out.

### Data sources

The PhysioNet Eye Tracking Dataset for the 12-Lead Electrocardiogram Interpretation of Medical Practitioners and Students, version 1.0.0 [20,21], comprises 63 readers interpreting ten ECGs: normal sinus rhythm, atrial fibrillation, hyperkalaemia, atrial flutter, ventricular tachycardia, Wolff-Parkinson-White syndrome, ventricular pacing, left bundle branch block, ST-elevation myocardial infarction and complete heart block. Viewing was recorded using Tobii Pro X2-60 at approximately 60 Hz [21]. Nine consultants formed one target population; the broader population contained these nine plus ten fellows and one resident (20 clinicians).

The local PTB-XL release was version 1.0.3 [22,23] and comprised 21,799 electrocardiograms. Fold 10 (test) contained 2,198 electrocardiograms from 1,904 patients and folds 1-9 (development) contained 19,601 from 16,965 patients, with no patient overlap; all 23 diagnostic subclasses were represented in fold 10, and 40 fold-10 electrocardiograms with no positive subclass label were retained as valid all-negative examples. Released labels were used without re-adjudication.

### Clinician gaze target construction

Gaze was assigned to twelve short diagnostic lead regions only; long rhythm strips, panels and any other areas not represented in the model input were excluded. Each reader’s per-ECG 12-lead dwell vector was normalised to sum to one, and vectors were averaged with equal reader weighting within each population (lead order I, II, III, aVR, aVL, aVF, V1–V6). The ten targets per population are shown in Supplementary Figures S1 and S2. Dwell describes viewing allocation, not the clinical reason for fixation: search difficulty, uncertainty and verification can all increase dwell [7,12].

### Image-derived electrocardiogram reconstruction

Because the gaze resource contains displayed images rather than digitised voltages, the twelve short lead traces were reconstructed into pseudo-waveforms. After locating short-lead rows and columns and excluding rhythm strips, each lead was resampled to 1,000 points and z-normalised. Ambiguous traces were checked visually and repaired with anchor-guided interpolation and local pixel snapping based only on trace quality. The final 10 × 12 × 1,000 × 1 array and lead order were frozen before training (checksum in Supplementary Table S1). These pseudo-waveforms are not equivalent to raw digital ECG voltage signals (Figure 2).

**Figure 2.**
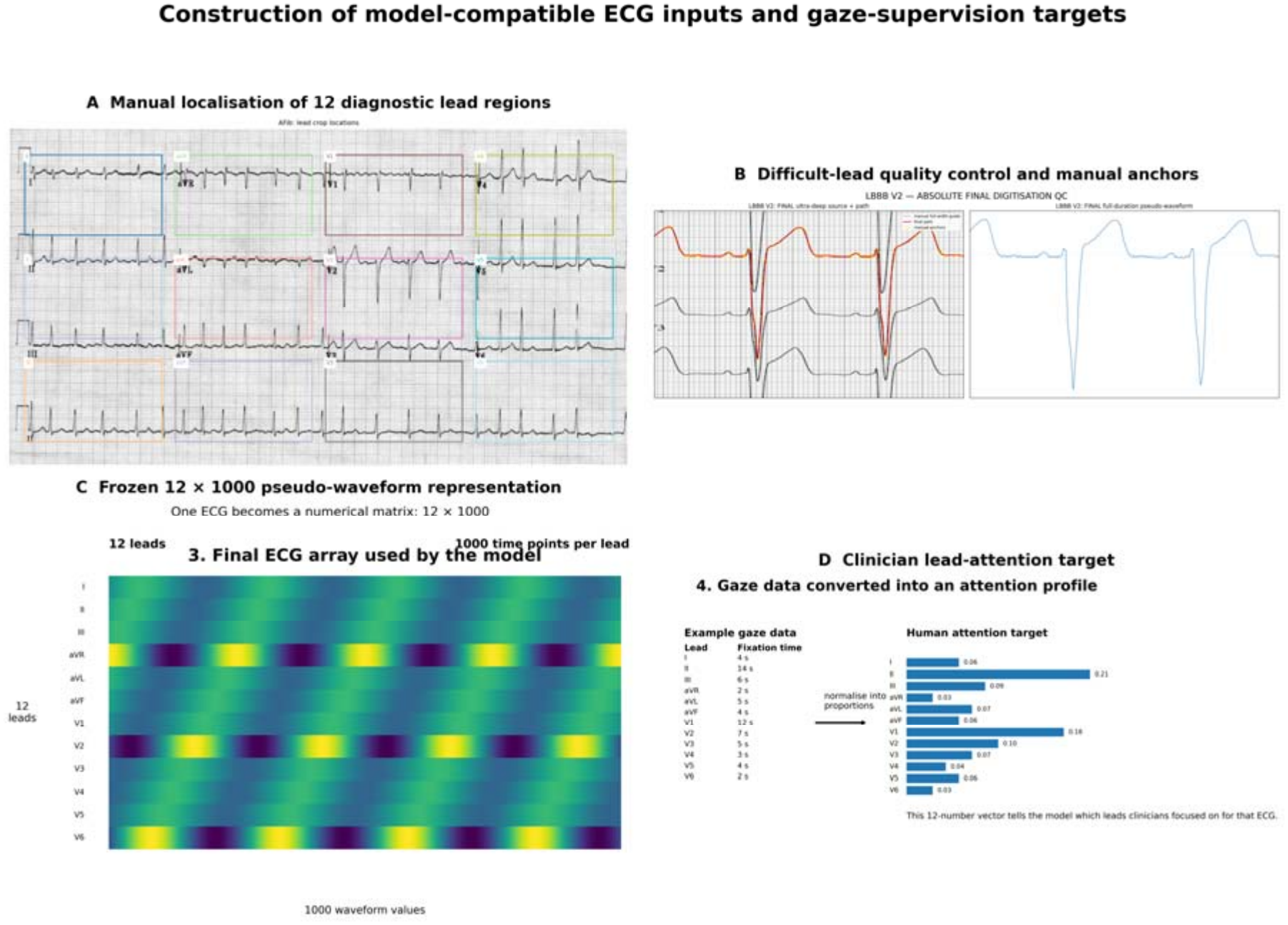
Construction of model-compatible electrocardiogram inputs and gaze targets. (A) Manual localisation of the 12 short diagnostic lead regions in the source image. (B) Difficult-lead quality control using manual anchors and overlay inspection. (C) Frozen 12 × 1,000 image-derived pseudo-waveform representation. (D) Example 12-lead clinician gaze distribution. All reported arms use the same corrected per-lead reconstruction, conventional fixed lead order, and frozen input tensor; superseded exploratory resize/lead-order variants are not part of this analysis. The reconstruction is not equivalent to original raw digital electrocardiogram voltage data.

### Pretrained electrocardiogram network

We used the publicly available residual 2D CNN of Elyamani et al. [5], pretrained on PTB-XL, with input 12 × 1,000 × 1 and 23 sigmoid diagnostic-subclass outputs (abbreviations in Table 1). Its weights served as the unchanged starting point for all arms. The same PTB-XL source dataset supplied fold-10 evaluation, so results represent source-domain held-out testing, not independent external validation [4,5,22,23].

### Activation-derived lead profile and functional check

The model-side measure was defined from layer add_3 (12 × 55 × 128), whose twelve-row spatial axis corresponds to the input leads. Root-mean-square activation was averaged across 55 temporal positions and 128 channels for each row, then normalised over the twelve rows. Convolutions preceding this readout retain the lead-row dimension; a later 12 × 1 convolution mixes leads downstream. This operational activation-derived lead profile is directly differentiable but is neither a formal attention mechanism nor a causal diagnostic attribution. Its relationship to lead contribution was tested independently through occlusion.

To examine whether the profile relates to the model’s functional use of each lead, a post hoc lead-occlusion analysis was performed with the unchanged base model on all 2,198 PTB-XL fold-10 electrocardiograms. Each z-normalised input lead was set to zero in turn, and functional lead influence was quantified as the mean absolute change in diagnostic logits across all 23 outputs and, separately, across positive labels only in records with at least one positive label. For each record, the Spearman correlation between the twelve-lead activation ranking and the occlusion-influence ranking was calculated, and agreement of the top-ranked lead was recorded (chance agreement 1/12, 8.3%).

### Gaze-supervised fine-tuning

The original experiment minimised mean squared error between the activation-derived 12-lead profile and the gaze target, without a diagnostic term. Fresh copies of the pretrained weights were used for each fold or arm, with only conv2d_10 trainable, BatchNorm parameters frozen and forward passes in inference mode. Training used Adam (learning rate 10□□), 25 epochs, batch size 16 and 40 Gaussian-noise copies per ECG (SD 0.03), identical across real-gaze and null-target arms. Augmentation increased optimisation samples, not independent ECGs.

### Question 1: target engagement

Five-fold ECG-level cross-validation withheld two complete ECGs per fold (seed 2026); models learned from eight independent gaze-labelled ECGs, and each held-out ECG represented a diagnosis absent from its fold’s gaze-training cases. The five held-out pairs were flutter/LBBB; STEMI/complete heart block; ventricular tachycardia/WPW; sinus rhythm/hyperkalaemia; and atrial fibrillation/ventricular pacing. No held-out gaze information entered the fold model. Agreement used cosine similarity, mean squared error and Jensen-Shannon divergence.

### Question 2: case specificity

A training-fold mean-gaze control averaged the twelve-lead gaze targets of the eight training electrocardiograms in each fold and compared this vector directly with each held-out target, without model training or access to the held-out case. A network that had learned case-specific gaze should agree with held-out gaze better than this population prior. Because gaze patterns are proportions summing to one, a centred cosine similarity, computed after subtracting the uniform pattern from both, was also reported, together with total-variation distance. At case level, the change in each held-out target’s dominant lead was compared with the shift required to reach the target.

### Question 3: functional preservation and attribution

For diagnostic evaluation, full-data real-gaze models and uniform and fixed permuted-target controls used all ten gaze-labelled ECGs and identical single-layer training conditions. Uniform targets placed 1/12 on each lead; the prespecified fixed derangement assigned every ECG another ECG’s gaze. Ten further unique derangements per population were frozen in a SHA-256 fingerprinted manifest before training. The unchanged base model and all gaze-only fine-tuned models (two real-gaze, one uniform and 22 permuted) each generated 23 sigmoid probabilities per PTB-XL fold-10 ECG (n = 2,198) and were assessed by macro area under the receiver operating characteristic curve (AUROC) and macro average precision (AP), without threshold tuning.

#### Post hoc joint diagnostic-plus-gaze experiment

To assess gaze supervision under a function-preserving regime, binary cross-entropy (BCE) on 19,601 PTB-XL development ECGs (folds 1 to 9) was combined with gaze-profile mean squared error (MSE) on the ten frozen gaze ECGs. Layers conv2d_10, conv2d_11 and three dense layers were trainable, with all BatchNorm layers frozen. Six arms shared identical PTB-XL batches and schedules: diagnostic-only, joint consultant, joint all-clinician, joint uniform, and two joint fixed-permutation controls. A single coefficient λ = 27.107 was fixed before training by equating initial diagnostic BCE (0.06043) to mean real-gaze MSE (0.002229); this equalises scalar loss magnitudes, not gradient influence. Adam learning rate was 10□ over four PTB-XL epochs (batches 128 diagnostic and 16 gaze; 24.64 effective passes through the 400 gaze augmentations). PTB-XL fold 10 was held out for all models. Gaze fit was assessed descriptively on the ten training ECGs only, as no independent gaze cases were available.

### Statistical analysis

Paired held-out ECG comparisons used two-sided Wilcoxon signed-rank tests; cosine, MSE and Jensen-Shannon divergence were interpreted as related measures without multiplicity adjustment. The gaze-only evaluation used 3,000 paired patient-cluster bootstrap draws, with 2,255 valid macro-metric samples because rare classes were absent in other resamples. Eleven gaze-derangement results per population were described without a formal permutation P value. The joint-loss experiment used the same patient-level bootstrap scheme (2,276 valid of 3,000). Absolute performance was rounded to three decimals in prose and four in tables, with additional precision retained for small contrasts.

### Supplementary representation-transfer analysis

A prespecified exploratory representation-transfer analysis on the independent Lobachevsky University ECG Database (LUDB; 200 records) [24] is reported in Supplementary Methods, Tables S2–S3 and Figure S3. Without matched null-target controls, it cannot support gaze-specific conclusions.

### Ethics, consent, and reproducibility

No new participants were recruited and no identifiable patient records were accessed. The eye-tracking study reported QBRI ethics approval (QBRI-IRB-2020-01-009) and written consent [20,21]. PTB-XL and LUDB are public de-identified datasets [22-24]. The cross-validation split, NumPy and TensorFlow randomisation used seed 2026; seed-derived generators were used for augmentation and derangements, with deterministic TensorFlow operations enabled where supported. Frozen source and weights checksums and the additional-permutation manifest hash are documented in Supplementary Table S1.

## Results

### Gaze-target audit and model-visible attention

Structural auditing confirmed twelve selected diagnostic lead regions for every reader and electrocardiogram, and all twenty clinicians in the combined target contributed positive viewing time within these regions for all ten electrocardiograms. Clinician attention was not confined to the short diagnostic leads represented in the model input: across the 200 clinician-by-electrocardiogram observations, the model-visible regions accounted for a mean of 63.5% of viewing time (median 65.4%, range 16.2% to 98.9%), leaving a mean of 36.5% directed elsewhere, predominantly at the rhythm strips.

### Question 1: target engagement

Gaze-only fine-tuning increased held-out cosine similarity (consultant 0.858 to 0.872; all clinicians 0.884 to 0.898; Wilcoxon P = 0.027 for both), with smaller mean squared error and Jensen-Shannon divergence (Table 2; Figure 3). Nine of ten ECGs improved on each metric; STEMI was the exception. Consultant and all-clinician gains were similar (difference in cosine gain 0.0008, P = 0.625), although ten cases cannot demonstrate equivalence. Audits confirmed that only conv2d_10 changed in the gaze-only arms.

**Table 2.** Held-out agreement between the activation-derived lead profile and clinician gaze, with the trainingfold mean-gaze control.

| **Population** | **Base profile cosine** | **Gaze-trained profile cosine** | **P, base vs gaze-trained** | **Training-fold mean-gaze cosine** | **Gaze-trained centred cosine** | **Mean-gaze centred cosine** | **P, gaze-trained vs mean gaze** |
|---|---|---|---|---|---|---|---|
| Consultants | 0.8584 | 0.8720 | 0.027 | 0.9294 | 0.0463 | 0.6775 | 0.002 |
| All clinicians | 0.8841 | 0.8984 | 0.027 | 0.9511 | 0.0598 | 0.7279 | 0.002 |

*P values are from two-sided paired Wilcoxon signed-rank tests of cosine similarity across ten held-out electrocardiograms; 0.002 is the smallest two-sided value attainable with ten pairs. Centred cosine subtracts the uniform 12-lead distribution from both vectors before cosine calculation. Mean squared error, Jensen-Shannon divergence, and total-variation distance likewise favoured the training-fold mean-gaze baseline. Agreement is with the prespecified activation proxy and does not establish causal diagnostic importance.*

**Figure 3.**
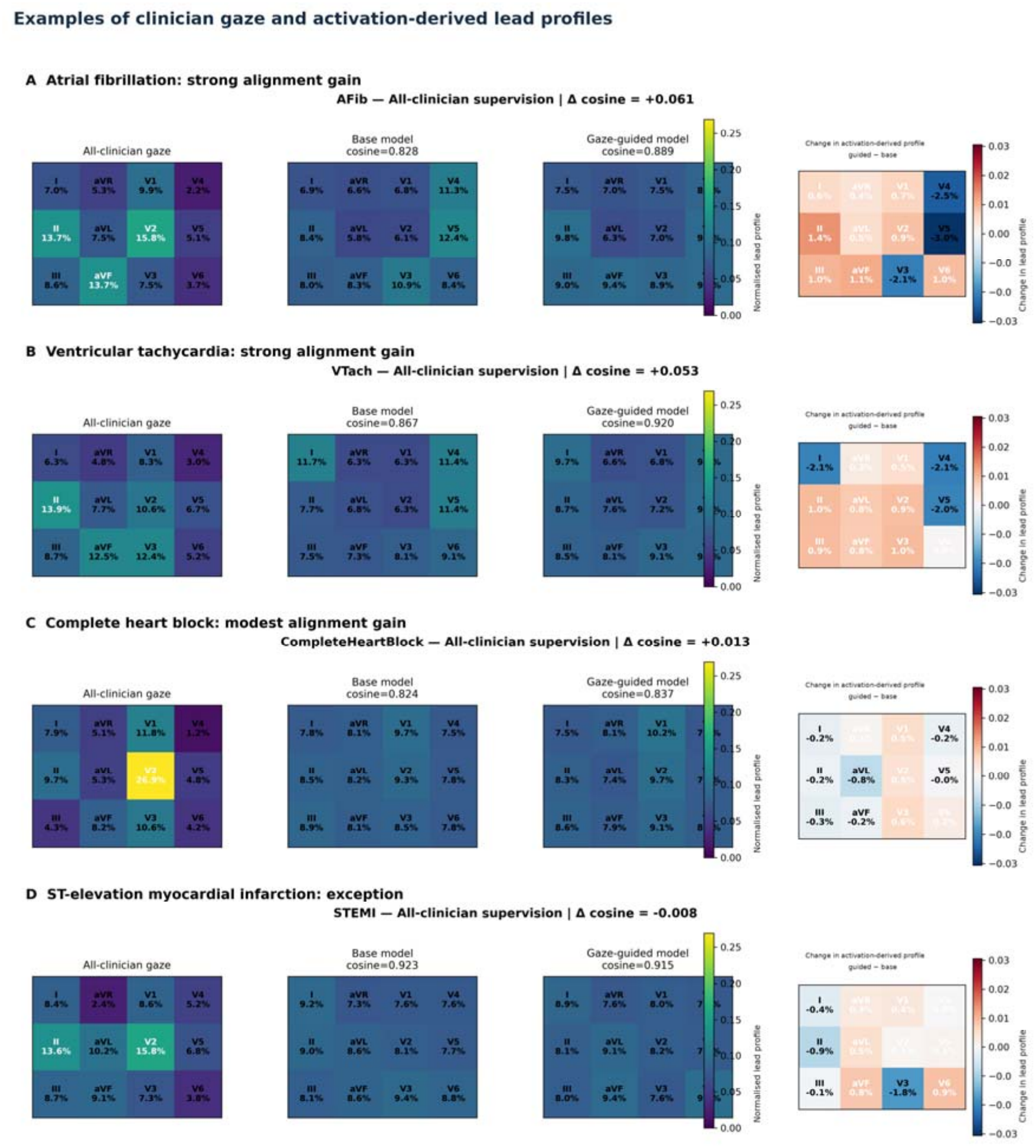
Illustrative full-data all-clinician activation-profile visualisations for atrial fibrillation, ventricular tachycardia, complete heart block, and ST-elevation myocardial infarction. Each row shows the clinician gaze target, the base activation-derived lead profile, the gaze-trained profile, and the gaze-trained-minus-base change. Complete heart block illustrates the distinction between global distributional similarity and case-specific target engagement: clinician gaze required a large increase in V2 emphasis, while the trained model barely shifted V2 from its baseline level. These are illustrative final-model visualisations; held-out comparisons are summarised in Table 2 and Figure 4. The activation-derived lead profile is an operational normalised RMS summary; visualisations do not establish causal lead attribution.

The activation-derived profile was weakly but predominantly positively related to functional lead influence. Across all 2,198 PTB-XL fold-10 electrocardiograms, the mean within-record Spearman correlation between the base model’s profile and all-output occlusion influence was 0.236 (SD 0.310; median 0.259, interquartile range 0.014 to 0.469) and was positive in 76.0% of records. For the 2,158 records with at least one positive label, the correlation with positive-label occlusion influence averaged 0.202 (SD 0.324; median 0.224, interquartile range −0.007 to 0.446), positive in 73.7%. The most active lead was also the most influential in 20.6% (all outputs) and 21.6% (positive-label outputs) of records, compared with 8.3% expected by chance.

### Question 2: case specificity

Case-specific gaze learning was not demonstrated. An untrained training-fold mean-gaze vector exceeded the gaze-trained model on held-out cosine similarity (consultants 0.929 vs 0.872; all clinicians 0.951 vs 0.898; paired P = 0.002 for both; Table 2, Figure 4). After removing the uniform component, centred cosine was 0.678 vs 0.046 and 0.728 vs 0.060, respectively. MSE, Jensen-Shannon divergence and total variation gave concordant findings. The dominant-lead case checks likewise showed minimal target-specific movement.

**Figure 4.**
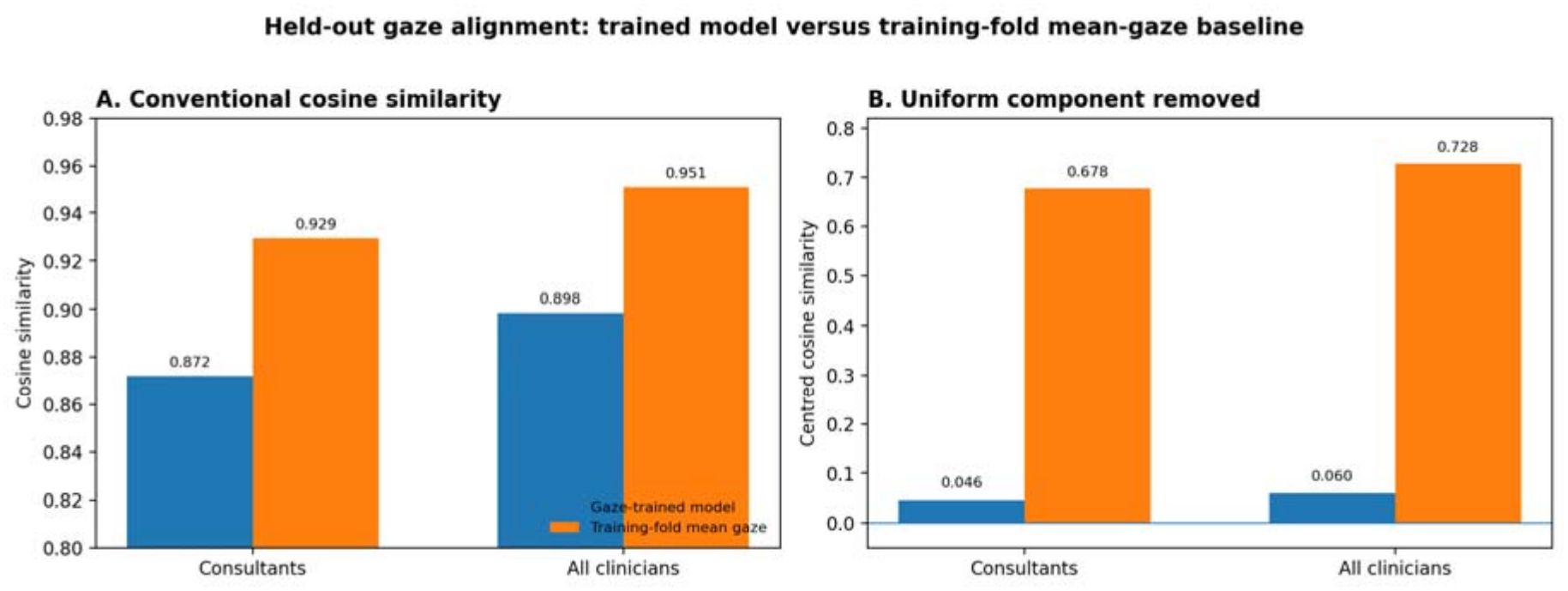
Held-out agreement between the activation-derived lead profile and clinician gaze, compared with a training-fold mean-gaze baseline. (A) Conventional cosine similarity for the gaze-trained profile and the non-trained training-fold mean-gaze vector. (B) Centred cosine similarity after subtracting the uniform 12-lead component. The mean-gaze baseline substantially outperformed the gaze-trained profile in both populations, demonstrating why improvement relative to an unchanged base model is insufficient to establish case-specific gaze learning.

Case-level analysis was concordant (Figure 3). For complete heart block, the consultant target placed 33.5% of model-visible gaze on V2, compared with 9.3% for the base profile; after fine-tuning, V2 emphasis remained 9.3%, approximately 0.2% of the required shift. The all-clinician target placed 26.9% of gaze on V2, with the same outcome, less than 0.4% of the required dominant-lead shift. More diffuse targets produced the largest cosine gains, showing that global agreement can improve without reproducing a target’s distinctive lead emphasis.

### Question 3: functional preservation and attribution

On PTB-XL fold 10, base macro AUROC/AP were 0.922/0.472 (Table 3, Figure 5). Gaze-only fine-tuning reduced AUROC to 0.893 for both target populations; the uniform and primary fixed-permutation controls fell to 0.899, 0.897 and 0.894, respectively. All five fine-tuned arms had lower discrimination than base, with patient-cluster bootstrap confidence intervals excluding zero for both macro metrics (Table 3).

**Table 3.**
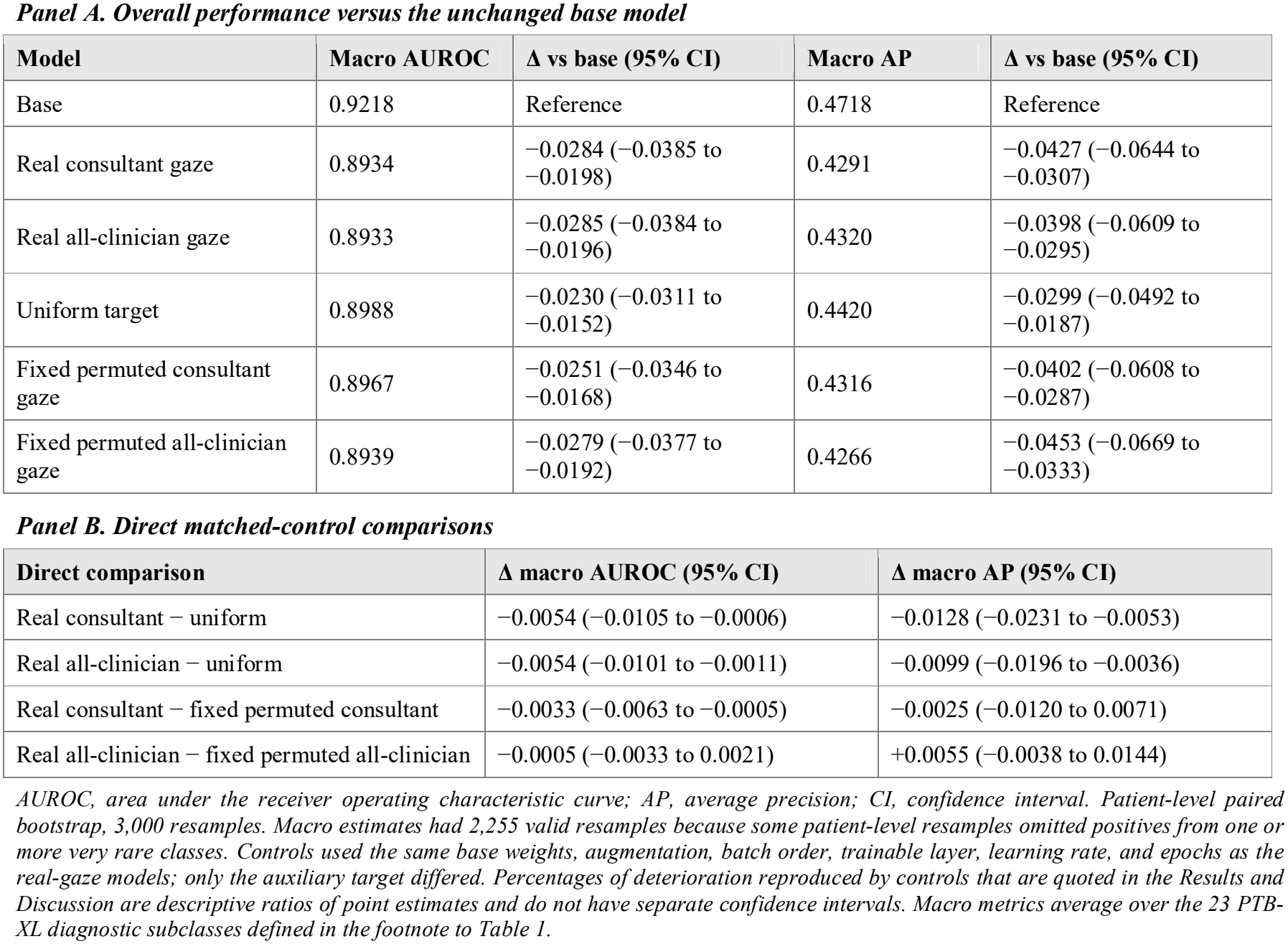
PTB-XL diagnostic performance after real and control auxiliary-target fine-tuning.

| ***Panel A. Overall performance versus the unchanged base model*** |  |  |  |  |
|---|---|---|---|---|
| **Model** | **Macro AUROC** | **Δ vs base (95% CI)** | **Macro AP** | **Δ vs base (95% CI)** |
| Base | 0.9218 | Reference | 0.4718 | Reference |
| Real consultant gaze | 0.8934 | −0.0284 (−0.0385 to −0.0198) | 0.4291 | −0.0427 (−0.0644 to −0.0307) |
| Real all-clinician gaze | 0.8933 | −0.0285 (−0.0384 to −0.0196) | 0.4320 | −0.0398 (−0.0609 to −0.0295) |
| Uniform target | 0.8988 | −0.0230 (−0.0311 to −0.0152) | 0.4420 | −0.0299 (−0.0492 to −0.0187) |
| Fixed permuted consultant gaze | 0.8967 | −0.0251 (−0.0346 to −0.0168) | 0.4316 | −0.0402 (−0.0608 to −0.0287) |
| Fixed permuted all-clinician gaze | 0.8939 | −0.0279 (−0.0377 to −0.0192) | 0.4266 | −0.0453 (−0.0669 to −0.0333) |
| ***Panel B. Direct matched-control comparisons*** |  |  |  |  |
| **Direct comparison** | **Δ macro AUROC (95% CI)** | **Δ macro AP (95% CI)** |  |  |
| Real consultant − uniform | −0.0054 (−0.0105 to −0.0006) | −0.0128 (−0.0231 to −0.0053) |  |  |
| Real all-clinician − uniform | −0.0054 (−0.0101 to −0.0011) | −0.0099 (−0.0196 to −0.0036) |  |  |
| Real consultant − fixed permuted consultant | −0.0033 (−0.0063 to −0.0005) | −0.0025 (−0.0120 to 0.0071) |  |  |
| Real all-clinician − fixed permuted all-clinician | −0.0005 (−0.0033 to 0.0021) | +0.0055 (−0.0038 to 0.0144) |  |  |

*AUROC, area under the receiver operating characteristic curve; AP, average precision; CI, confidence interval. Patient-level paired bootstrap, 3,000 resamples. Macro estimates had 2,255 valid resamples because some patient-level resamples omitted positives from one or more very rare classes. Controls used the same base weights, augmentation, batch order, trainable layer, learning rate, and epochs as the real-gaze models; only the auxiliary target differed. Percentages of deterioration reproduced by controls that are quoted in the Results and Discussion are descriptive ratios of point estimates and do not have separate confidence intervals. Macro metrics average over the 23 PTB-XL diagnostic subclasses defined in the footnote to Table 1.*

**Figure 5.**
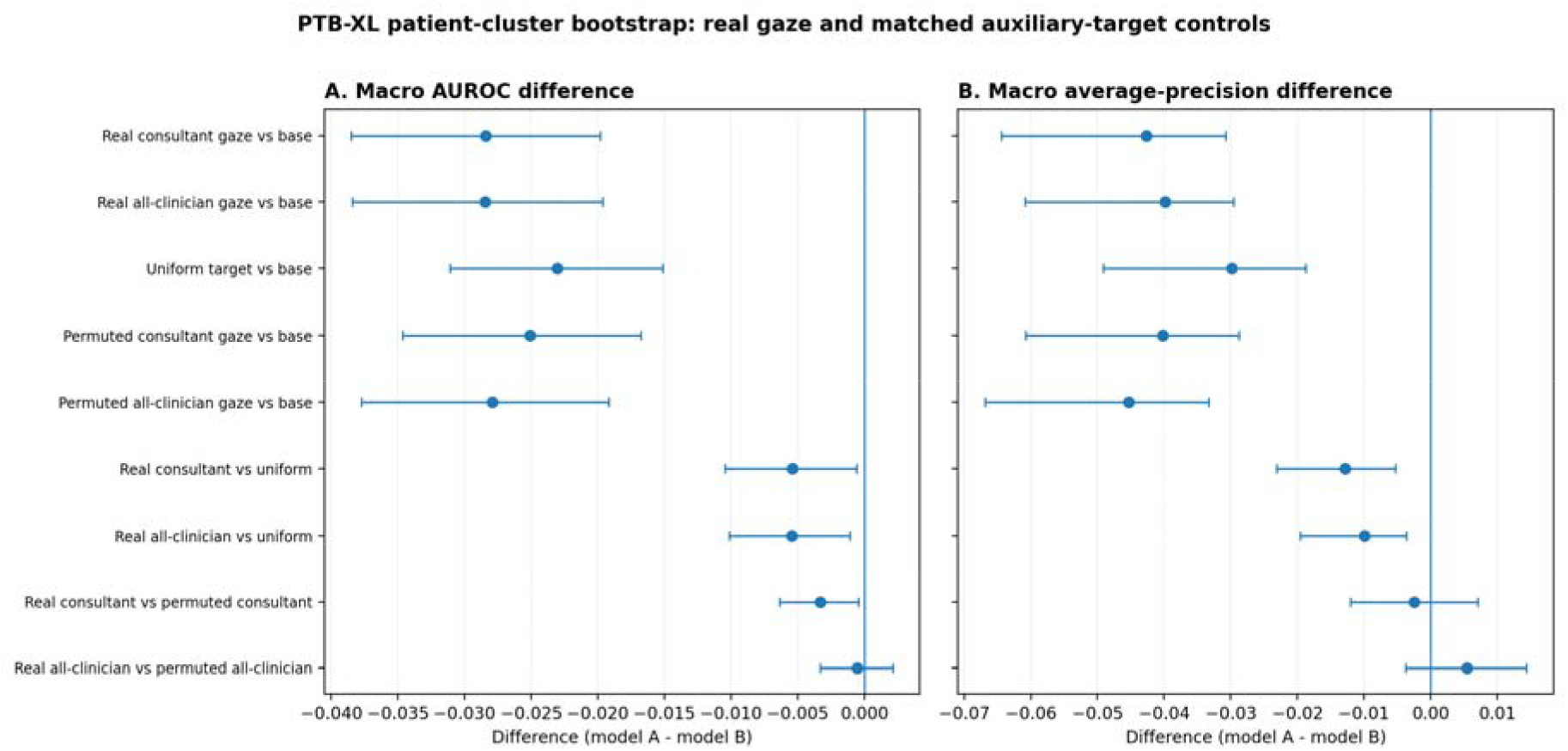
PTB-XL patient-cluster bootstrap comparison of real gaze and matched auxiliary-target controls. Points show macro-metric differences and horizontal bars show percentile 95% confidence intervals for the primary fixed controls. (A) Macro area-under-the-receiver-operating-characteristic-curve differences. (B) Macro average-precision differences. Uniform and permuted targets reproduced most of the gaze-versus-base deterioration. Real consultant gaze showed a small additional area-under-the-curve decrement relative to the fixed permuted consultant control, with its confidence interval excluding zero, while the corresponding average-precision interval crossed zero. Real all-clinician gaze did not clearly differ from its permuted control on either macro metric. Ten additional derangements per population (Table 4) showed that correctly matched gaze lay within the range of permuted targets.

In direct matched comparisons (Table 3, Panel B), real gaze was slightly worse than the uniform target (macro AUROC difference −0.0054, 95% CI −0.0105 to −0.0006, for consultants; −0.0054, −0.0101 to −0.0011, for all clinicians). Against the fixed permuted control, consultant gaze showed a small additional macro AUROC decrement (−0.0033, −0.0063 to −0.0005), with a macro AP difference crossing zero (−0.0025, −0.0120 to 0.0071); all-clinician gaze did not differ clearly on either metric (macro AUROC −0.0005, −0.0033 to 0.0021; macro AP +0.0055, −0.0038 to 0.0144).

**Table 4.**
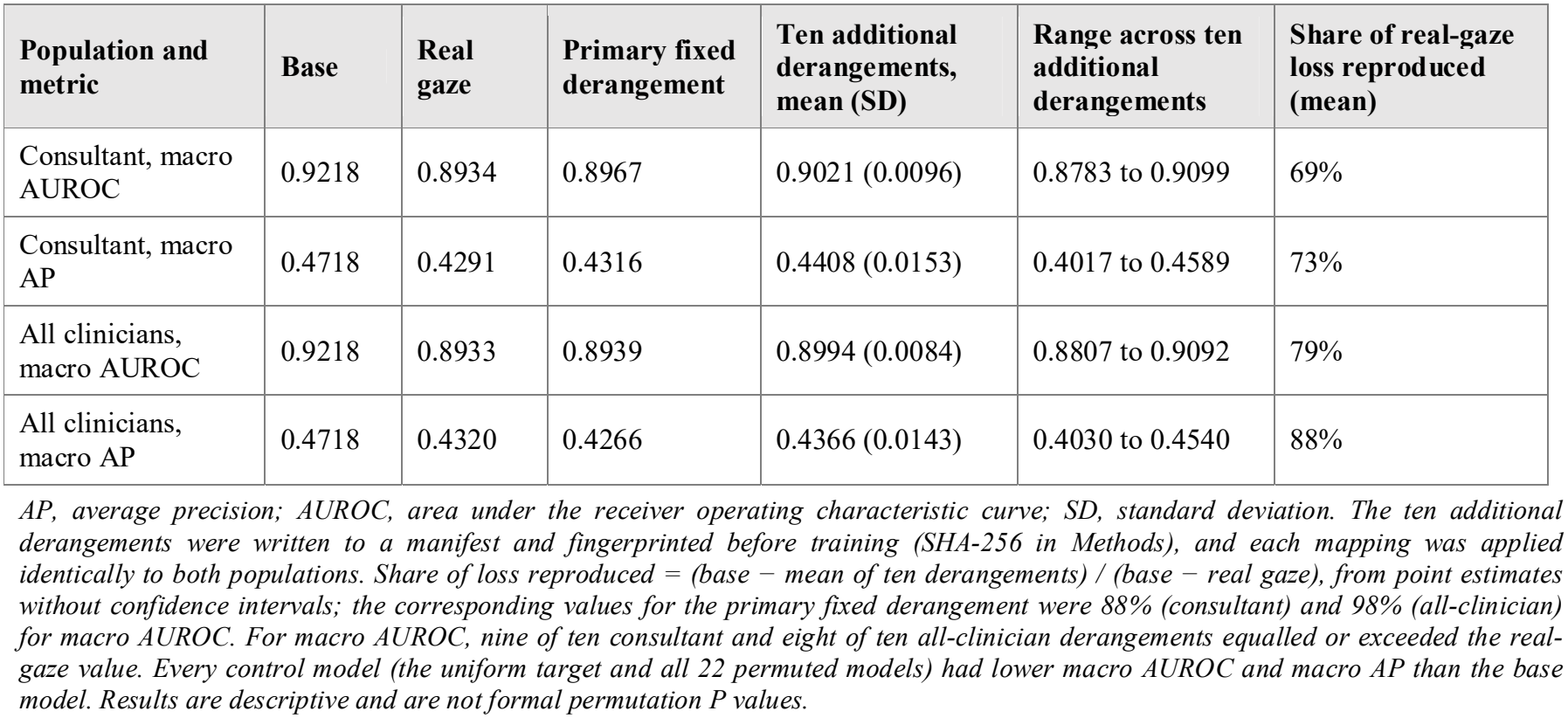
Sensitivity of diagnostic attribution to the choice of gaze derangement.

| **Population and metric** | **Base** | **Real gaze** | **Primary fixed derangement** | **Ten additional derangements, mean (SD)** | **Range across ten additional derangements** | **Share of real-gaze loss reproduced (mean)** |
|---|---|---|---|---|---|---|
| Consultant, macro AUROC | 0.9218 | 0.8934 | 0.8967 | 0.9021 (0.0096) | 0.8783 to 0.9099 | 69% |
| Consultant, macro AP | 0.4718 | 0.4291 | 0.4316 | 0.4408 (0.0153) | 0.4017 to 0.4589 | 73% |
| All clinicians, macro AUROC | 0.9218 | 0.8933 | 0.8939 | 0.8994 (0.0084) | 0.8807 to 0.9092 | 79% |
| All clinicians, macro AP | 0.4718 | 0.4320 | 0.4266 | 0.4366 (0.0143) | 0.4030 to 0.4540 | 88% |

*AP, average precision; AUROC, area under the receiver operating characteristic curve; SD, standard deviation. The ten additional derangements were written to a manifest and fingerprinted before training (SHA-256 in Methods), and each mapping was applied identically to both populations. Share of loss reproduced = (base − mean of ten derangements) / (base − real gaze), from point estimates without confidence intervals; the corresponding values for the primary fixed derangement were 88% (consultant) and 98% (all-clinician) for macro AUROC. For macro AUROC, nine of ten consultant and eight of ten all-clinician derangements equalled or exceeded the real-gaze value. Every control model (the uniform target and all 22 permuted models) had lower macro AUROC and macro AP than the base model. Results are descriptive and are not formal permutation P values.*

To assess fixed-shuffle sensitivity, ten additional derangements were applied to each population (Table 4). All 23 null-target models (uniform plus 22 permutations) reduced macro AUROC versus base (range 0.878–0.910). Across the ten new derangements, permuted macro AUROC averaged 0.902 (SD 0.0096) and 0.899 (SD 0.0084) for consultant and all-clinician targets; matched gaze was 0.893 in each. Nine of ten consultant and eight of ten all-clinician derangements performed at least as well as the corresponding real-gaze model. Mean mismatched controls reproduced 69% and 79% of the real-gaze AUROC loss, versus 88% and 98% for the original fixed shuffle; these are descriptive ratios without uncertainty intervals.

#### Post hoc joint diagnostic-plus-gaze experiment

With diagnostic supervision retained, fold-10 discrimination was preserved in every arm (Table 5). Macro AUROC was 0.923 and macro AP 0.475 after diagnostic-only, consultant-gaze and all-clinician-gaze fine-tuning, and the uniform and permuted controls were within 0.0001 of these values. Versus diagnostic-only, paired macro AUROC differences were −0.000016 (95% CI −0.000400 to 0.000354) for consultant gaze and −0.000001 (−0.000415 to 0.000398) for all-clinician gaze (2,276 valid patient-cluster bootstrap replicates; Supplementary Tables S4 and S5). However, real-gaze targets were barely fitted, even on the ten training ECGs: cosine similarity moved from 0.858 to 0.860 (consultant) and from 0.884 to 0.886 (all clinicians), and fit to permuted targets was of similar magnitude (0.861 and 0.883). By contrast, the uniform arm fitted its target closely (cosine 0.989) without loss of discrimination (Supplementary Table S6). The joint regime therefore preserved diagnostic function, but the gaze term was not meaningfully engaged, so this experiment could not test whether correctly matched gaze adds diagnostic value.

**Table 5.** Post hoc joint diagnostic-plus-gaze fine-tuning: PTB-XL fold-10 discrimination and training-ECG gaze fit.

| **Arm** | **Macro AUROC** | **Macro AP** | **Δ macro AUROC vs diagnostic-only (95% CI)** | **Training-ECG cosine with own target** |
|---|---|---|---|---|
| Base pretrained | 0.9218 | 0.4718 | −0.001197 (−0.002661 to 0.000178) | 0.8584 (consultant); 0.8841 (all clinicians) |
| Diagnostic-only | 0.9230 | 0.4746 | Reference | 0.8584 (consultant); 0.8841 (all clinicians) |
| Joint consultant gaze | 0.9230 | 0.4746 | −0.000016 (−0.000400 to 0.000354) | 0.8602 |
| Joint all-clinician gaze | 0.9230 | 0.4747 | −0.000001 (−0.000415 to 0.000398) | 0.8860 |
| Joint uniform target | 0.9229 | 0.4745 | −0.000123 (−0.000402 to 0.000156) | 0.9887 |
| Joint fixed permuted consultant gaze | 0.9230 | 0.4747 | −0.000041 (−0.000463 to 0.000359) | 0.8607 |
| Joint fixed permuted all-clinician gaze | 0.9229 | 0.4747 | −0.000089 (−0.000555 to 0.000360) | 0.8829 |

*AP, average precision; AUROC, area under the receiver operating characteristic curve; CI, confidence interval. All arms started from the same pretrained weights and shared identical PTB-XL minibatches, schedules and trainable layers (conv2d\_10, conv2d\_11 and three dense layers); only the auxiliary gaze target differed, and the diagnostic-only arm had no gaze term. Differences are paired patient-cluster bootstrap estimates (2,276 valid of 3,000 replicates), given to six decimal places because they are very small; the base row shows base minus diagnostic-only. Training-ECG cosine is the in-sample cosine similarity between each model’s activation-derived lead profile and its own auxiliary target on the ten gaze ECGs (for base and diagnostic-only, with the consultant and all-clinician targets); it does not measure held-out gaze learning. Real-minus-permuted macro AUROC differences were +0.000024 (−0.000067 to 0.000128) for consultant and +0.000088 (−0.000022 to 0.000218) for all-clinician gaze. Full results, including micro-averaged metrics and AP contrasts, are in Supplementary Tables S4 to S6.*

### Supplementary representation-transfer findings

Exploratory LUDB findings were task-dependent, with lower ischaemia-related discrimination after gaze supervision and no clear difference for conduction abnormality or left ventricular hypertrophy (Supplementary Table S2); without matched controls, they are not interpreted as gaze-specific.

## Discussion

### Principal findings

Three questions were separated. Gaze-only optimisation modestly increased target agreement, yet did not outperform an untrained training-fold mean-gaze prior. Source-domain discrimination deteriorated, but the loss was shared by uniform and permuted controls. A post hoc joint diagnostic-plus-gaze experiment retained discrimination at the diagnostic-only level, but real-gaze targets were barely fitted under that regime, so it could not test whether gaze adds diagnostic value. Thus neither target engagement nor a change in diagnostic function should be attributed to specific human viewing behaviour without matched controls.

### Interpreting apparent gaze alignment

Movement toward a target must be distinguished from learning target-specific information. Because gaze distributions are non-negative and sum to one, cosine similarity was high before training (0.858 and 0.884) and can remain high when the most distinctive lead differs; removing the shared uniform component made the gap between the trained network and the mean-gaze prior far clearer. Complete heart block showed that a target’s distinctive lead emphasis can remain unlearned while global agreement improves.

The negative case-specificity result concerns an extremely small experiment. Each fold used only eight independent gaze-labelled ECGs, and each held-out case represented an unseen diagnosis; noise augmentation added no new cases. Training modified one layer with a small learning rate, and the model lacked the clinical context known to influence human interpretation [10,25]. More powerful training on larger gaze datasets may behave differently.

Neither dwell nor activation should be equated with diagnostic importance. The occlusion analysis indicates that the activation-derived profile is related to the model’s functional use of the leads, but only weakly; it is best read as an operational, representation-level summary, and its agreement with clinician gaze as target engagement only.

### Interpreting the loss of diagnostic performance

A gaze-only objective does not explicitly maintain classification, while downstream layers were frozen and could not adjust to any perturbation of conv2d_10. This provides a plausible explanation for the diagnostic deterioration observed under all auxiliary targets. Between-target variability was substantial (permuted macro AUROC 0.878 to 0.910). The joint-loss experiment was performed separately, with diagnostic supervision and an adaptable trainable tail. It maintained source-domain classification across all matched arms, including a uniform arm that fitted its target closely (training-ECG cosine 0.989), which suggests that, once diagnostic supervision is retained and downstream layers can adapt, an auxiliary target need not cost discrimination. Because the two regimes differed in objective, data and trainable layers, this protection cannot be attributed to any single change.

Matched gaze fell within the permutation range, and the distribution across alternative shuffles was wider than the small fixed-permutation consultant contrast. A gaze-specific performance penalty cannot be established from that contrast. The repeated permutations are descriptive sensitivity analyses, not an exact permutation test.

Under joint training, the gaze term was barely engaged: training-ECG cosine with real-gaze targets changed by less than 0.002, a similar degree of fit to that achieved for permuted targets, whereas a uniform target was fitted closely. A target that is identical for every ECG can be met by a global change in the representation, whereas case-varying gaze requires case-specific adjustment, echoing the case-specificity result of the gaze-only experiment. The absence of a gaze-specific diagnostic difference is therefore expected and does not show that gaze is unhelpful; the experiment establishes preservation of function, not the value of gaze. The fixed λ matched *initial loss magnitudes*, not gradient contributions, and only ten gaze-labelled cases were available for the auxiliary task. Testing gaze as an auxiliary signal will require a larger gaze dataset, a weighting shown to engage the gaze target, and independent gaze evaluation.

### Consultant versus broader clinician gaze

The two reader populations produced similar held-out gaze-only target engagement and classification losses. A small consultant difference against one fixed shuffle was not robust to alternative derangements and should not be interpreted as expertise-specific. With only ten ECGs and nine consultants, reader-expertise interactions cannot be inferred.

### Implications for gaze-supervised medical artificial intelligence

The study suggests a practical three-step validation sequence for medical gaze supervision: demonstrate target engagement relative to an unchanged model; establish specificity against training-fold mean, uniform and several permuted gaze targets; then evaluate diagnostic preservation against matched diagnostic-only training on held-out data. Independent checks of the model-side measure, such as lead occlusion, further restrict interpretability claims. Comparable safeguards are relevant wherever human viewing is used for model supervision [13-19].

### Strengths and limitations

Strengths include explicit separation of the three questions, electrocardiogram-level cross-validation, identical frozen inputs and schedules across arms, weight audits, a functional check of the model-side measure, eleven frozen permutations per population, and patient-cluster bootstrap inference.

Limitations include only ten independent gaze-labelled ECGs, each a different diagnosis; omission of temporal scan paths and 36.5% of viewing outside model-visible short leads; image-derived rather than digital waveform inputs; and source-domain PTB-XL evaluation without label re-adjudication. The add_3 RMS profile remains an incomplete attribution proxy, and zeroing leads may generate out-of-distribution inputs. All controls and the joint experiment were added post hoc. In the joint experiment, gaze fit was assessed only on the ten training ECGs, real-gaze targets were barely fitted, and a single fixed shuffle was used for the permuted controls.

All experiments used one pretrained backbone. The unchanged model’s macro AUROC (0.922) falls within the 0.908–0.943 range reported for models on the same 23-subclass PTB-XL task in the Al-Masud et al. benchmark [26], although splits and preprocessing limit direct comparison. Later 12×1 convolution in this architecture combines leads, whereas other modern backbones mix lead channels at input; applying a similar lead-profile objective to them would require a newly validated readout. Joint-loss weights were fixed by initial scalar loss magnitude rather than tuned for clinical outcomes; λ should not be interpreted as balancing gradients. The absence of independent gaze cases and clinical context limits generalisability.

## Conclusion

In this ten-ECG matched-control proof-of-concept, gaze-only fine-tuning moved an activation-derived lead profile toward clinician viewing but did not establish case-specific gaze learning beyond a training-fold mean-gaze prior. Uniform and mismatched gaze reproduced the diagnostic deterioration, so the loss was not specific to correctly matched gaze. In a separate post hoc experiment, joint diagnostic-and-gaze training preserved PTB-XL discrimination, but real-gaze targets were barely fitted, so the experiment could not establish whether gaze adds diagnostic value. The study therefore shows neither that clinician gaze is useful nor that it is harmful. Claims about gaze supervision require population-mean and shuffled-target controls, checks on functional attribution, comparison with diagnostic-only training, and evidence that the gaze target was actually engaged.

## Data Availability

All datasets analysed in this study are publicly available through PhysioNet: the Eye Tracking Dataset for the 12-Lead Electrocardiogram Interpretation of Medical Practitioners and Students (version 1.0.0; https://doi.org/10.13026/gsr5-8b11), PTB-XL (version 1.0.3; https://doi.org/10.13026/kfzx-aw45), and the Lobachevsky University Electrocardiography Database (LUDB; https://physionet.org/content/ludb/1.0.1/).
The principal results and summary data are presented in the manuscript and supplementary materials. Additional derived data generated during the analyses are available from the corresponding author upon reasonable request, subject to applicable data-use conditions. The analysis code and experimental configuration files are publicly available at https://github.com/Anurup97/ecg-gaze-supervision/releases/tag/v1.0.0.

## Acknowledgements

The authors thank Mary Price, Evidence Specialist, Maidstone and Tunbridge Wells NHS Trust Library and Knowledge Service, for undertaking structured evidence searches that informed the literature review and contextualisation of this study. The searches addressed: clinical context and gaze behaviour during electrocardiogram interpretation (3 July 2026); expert and novice electrocardiogram gaze patterns (10 July 2026); eye-tracking research methods (16 July 2026); use of clinician gaze to train artificial-intelligence models in medical imaging (3 August 2026); electrocardiogram artificial-intelligence architecture and explainability approaches (4 August 2026); and systematic-review evidence concerning artificial intelligence in electrocardiogram interpretation (6 August 2026).

## Use of artificial intelligence tools

Generative artificial-intelligence tools were used for code troubleshooting, figure development, language editing, and manuscript drafting support. They were also used to clean and organise the analysis code and accompanying documentation before upload to a public GitHub repository. All analysis definitions, numerical outputs, source citations, interpretations, and final manuscript content were reviewed by the authors. No artificial-intelligence system is listed as an author.

## Consent

No identifiable patient information is presented. The original eye-tracking study obtained participant written informed consent as described in the source publication and dataset documentation [20,21]. No new consent was sought for this secondary analysis of public de-identified datasets.

## Data and Code availability

The datasets are publicly available: PhysioNet Eye Tracking Dataset version 1.0.0 [21], PTB-XL version 1.0.3 [23], and LUDB [24]. Analysis code, including the permutation and joint-loss manifests, is publicly available in the study’s GitHub repository (https://github.com/Anurup97/ecg-gaze-supervision/releases/tag/v1.0.0). Derived output tables not included in the repository are available from the corresponding author on reasonable request. SHA-256 fingerprints of the frozen inputs, models and manifests are listed in Supplementary Table S1 so that shared files can be verified against those analysed. Dataset licences and patient privacy restrictions apply to redistribution of raw source records.

## References

1. Kilic ME, Arayici ME, Akbilgic O, Yilmaz MB. Diagnostic accuracy of machine learning algorithms in electrocardiogram-based heart failure detection: a systematic review and meta-analysis. Can J Cardiol 2026;42:824–839. doi:10.1016/j.cjca.2025.12.022.

2. Lee Y, Ahn JH, Yang HM. Artificial intelligence-enhanced electrocardiography for acute myocardial infarction detection: a systematic review. Cardiovasc Diagn Ther 2026;16. doi:10.21037/cdt-2025-aw-561.

3. Taleban A, Sparapani R, Noffke P, Zlochiver S, Lu Q, Widlansky ME, et al. Explainable artificial intelligence in electrocardiography: a systematic review. Biomed Signal Process Control 2026;114:109325. doi:10.1016/j.bspc.2025.109325.

4. Strodthoff N, Wagner P, Schaeffter T, Samek W. Deep learning for ECG analysis: benchmarks and insights from PTB-XL. IEEE J Biomed Health Inform 2021;25:1519–1528. doi:10.1109/JBHI.2020.3022989.

5. Elyamani HA, Salem MA, Melgani F, Yhiea NM. Deep residual 2D convolutional neural network for cardiovascular disease classification. Sci Rep 2024;14:22040. doi:10.1038/s41598-024-72382-3.

6. Amani-Beni R, Darouei B, Kakavand N, Eshraghi R, Seifnezhad A, Movahed MR, et al. A systematic review of eye-tracking technology in electrocardiogram interpretation research. Eur J Med Res 2026;31:50. doi:10.1186/s40001-025-03635-8.

7. Breen CJ, Bond R, Finlay D. An evaluation of eye tracking technology in the assessment of 12 lead electrocardiography interpretation. J Electrocardiol 2014;47:922–929. doi:10.1016/j.jelectrocard.2014.08.008.

8. Wood G, Batt J, Appelboam A, Harris A, Wilson MR. Exploring the impact of expertise, clinical history, and visual search on electrocardiogram interpretation. Med Decis Making 2014;34:75–83. doi:10.1177/0272989X13492016.

9. Davies A, Harper S, Vigo M, Jay C. Using simultaneous scanpath visualization to investigate the relationship between accuracy and eye movement during medical image interpretation. J Eye Mov Res 2018;10(5). doi:10.16910/jemr.10.5.11.

10. Davies A, Harper S, Vigo M, Jay C. Investigating the effect of clinical history before electrocardiogram interpretation on the visual behavior and interpretation accuracy of clinicians. Sci Rep 2019;9:11300. doi:10.1038/s41598-019-47830-0.

11. Attar ET. Eye-tracking biomarkers of clinical expertise in ECG interpretation: statistical and machine learning evidence. Front Med (Lausanne) 2026;12:1704829. doi:10.3389/fmed.2025.1704829.

12. DiGirolamo GJ, Kakouros N, Sorcini F, Sorcini SP, Dupell J, Sands EK, et al. Non-conscious detection of ST-segment elevation during physician ECG interpretation. J Electrocardiol 2025;93:154148. doi:10.1016/j.jelectrocard.2025.154148.

13. Wang S, Ouyang X, Liu T, Wang Q, Shen D. Follow My Eye: using gaze to supervise computer-aided diagnosis. IEEE Trans Med Imaging 2022;41:1688–1698. doi:10.1109/TMI.2022.3146973.

14. Neves J, Hsieh C, Nobre IB, Sousa SC, Ouyang C, Maciel A, et al. Shedding light on AI in radiology: a systematic review and taxonomy of eye gaze-driven interpretability in deep learning. Eur J Radiol 2024;172:111341. doi:10.1016/j.ejrad.2024.111341.

15. Duan J, Zhang M, Song M, Xu X, Lu H. Eye tracking-enhanced deep learning for medical image analysis: a systematic review on data efficiency, interpretability, and multimodal integration. Bioengineering (Basel) 2025;12:954. doi:10.3390/bioengineering12090954.

16. Ghezloo F, Chang OH, Knezevich SR, Shaw KC, Thigpen KG, Reisch LM, et al. Robust ROI detection in whole slide images guided by pathologists’ viewing patterns. J Imaging Inform Med 2025;38:439–454. doi:10.1007/s10278-024-01202-x.

17. Guo Z, Hu Y, Ge P, Chan IN, Yan T, Wong PK, et al. Enhancing colorectal polyp classification using gaze-based attention networks. PeerJ Comput Sci 2025;11:e2780. doi:10.7717/peerj-cs.2780.

18. Nan T, Zheng S, Qiao S, Quan H, Gao X, Niu J, et al. Deep learning quantifies pathologists’ visual patterns for whole slide image diagnosis. Nat Commun 2025;16:5493. doi:10.1038/s41467-025-60307-1.

19. Chen J, Duan H, Zhang X, Gao B, Grau V, Han J. From gaze to insight: bridging human visual attention and vision language model explanation for weakly-supervised medical image segmentation. IEEE Trans Med Imaging 2026;45:1051–1061. doi:10.1109/TMI.2025.3616598.

20. Tahri Sqalli M, Al-Thani D, Elshazly MB, Al-Hijji M, Alahmadi A, Sqalli Houssaini Y. Understanding cardiology practitioners’ interpretations of electrocardiograms: an eye-tracking study. JMIR Hum Factors 2022;9:e34058. doi:10.2196/34058.

21. [dataset] Tahri Sqalli M, Al-Thani D, Elshazly M, Al-Hijji M. Eye Tracking Dataset for the 12-Lead Electrocardiogram Interpretation of Medical Practitioners and Students. Version 1.0.0. PhysioNet; 2022. doi:10.13026/gsr5-8b11.

22. Wagner P, Strodthoff N, Bousseljot RD, Kreiseler D, Lunze FI, Samek W, et al. PTB-XL, a large publicly available electrocardiography dataset. Sci Data 2020;7:154. doi:10.1038/s41597-020-0495-6.

23. [dataset] Wagner P, Strodthoff N, Bousseljot R-D, Samek W, Schaeffter T. PTB-XL, a large publicly available electrocardiography dataset. Version 1.0.3. PhysioNet; 2022. doi:10.13026/kfzx-aw45.

24. Kalyakulina AI, Yusipov II, Moskalenko VA, Nikolskiy AV, Kozlov AA, Zolotykh NY, et al. LUDB: a new open-access validation tool for electrocardiogram delineation algorithms. IEEE Access 2020;8:186181–186190. doi:10.1109/ACCESS.2020.3029211.

25. Viljoen CA, Millar RS, Manning K, Hoevelmann J, Burch VC. Clinically contextualised ECG interpretation: the impact of prior clinical exposure and case vignettes on ECG diagnostic accuracy. BMC Med Educ 2021;21:417. doi:10.1186/s12909-021-02854-x.

26. Al-Masud MA, Lopez Alcaraz JM, Strodthoff N. Benchmarking ECG FMs: a reality check across clinical tasks. In: International Conference on Learning Representations (ICLR) 2026. p. 146689–146715. arXiv:2509.25095.

